# Unsupervised clustering of missense variants in the *HNF1A* gene using multidimensional functional data aids clinical interpretation

**DOI:** 10.1101/19010900

**Authors:** Sara Althari, Laeya A. Najmi, Amanda J. Bennett, Ingvild Aukrust, Jana K. Rundle, Kevin Colclough, Janne Molnes, Alba Kaci, Sameena Nawaz, Timme van der Lugt, Neelam Hassanali, Anders Molven, Sian Ellard, Mark I. McCarthy, Lise Bjørkhaug, Pål Rasmus Njølstad, Anna L. Gloyn

## Abstract

**Background:** Exome sequencing in diabetes presents a diagnostic challenge as depending on frequency, functional impact and genomic and environmental contexts, *HNF1A* variants can cause Maturity-onset Diabetes of the Young (MODY), increase type 2 diabetes risk, or be benign. A correct diagnosis matters as it informs on treatment, progression, and family risk. We describe a multi-dimensional functional dataset of 73 *HNF1A* missense variants identified in exomes of 12,940 individuals. Our aim was to develop an analytical framework for stratifying variants along the *HNF1A* phenotypic continuum to facilitate diagnostic interpretation.

**Methods:** *HNF1A* variant function was determined by 4 different molecular assays. Structure of the multi-dimensional dataset was explored using principal component analysis, k-means, and hierarchical clustering. Weights for tissue-specific isoform expression and functional domain were integrated. Functionally annotated variant subgroups were used to re-evaluate genetic diagnoses in national MODY diagnostic registries.

**Findings:** *HNF1A* variants demonstrated a range of behaviours across the assays. The structure of the multi-parametric data was shaped primarily by transactivation. Using unsupervised learning methods, we obtained high-resolution functional clusters of the variants which separated known causal MODY variants from benign and type 2 diabetes risk variants and led to reclassification of 4% and 9% of *HNF1A* variants identified in the UK and Norway MODY diagnostic registries, respectively.

**Interpretation:** Our proof-of-principle analyses facilitated informative stratification of *HNF1A* variants along the continuum, allowing improved evaluation of clinical significance, management and precision medicine in diabetes clinics. Transcriptional activity appears a superior readout supporting pursuit of transactivation-centric experimental designs for high-throughput functional screens.

**Funding:** Wellcome Trust, National Institute for Health Research (NIHR) Oxford Biomedical Research Centre (BRC), European Research Council, Norwegian Research Council, Stiftelsen Kristian Gerhard Jebsen, Western Norway Regional Health Authority, Novo Nordisk Fonden, Royal Norwegian Diabetes Foundation.

**Research in context:** *Evidence before the study:* Molecular characterisation pipelines for studying the function of transcription factors consist primarily of *in vitro* cellular assays which interrogate transcriptional activity, protein abundance, localisation of the transcription factor to the nucleus, and binding to relevant DNA recognition sequences. The experimental techniques used to explore these mechanisms *in vitro* vary in robustness and reliability. There exist a wide variety of reported functional consequences of *HNF1A* variants in the literature, a gene causing the most common form of Maturity-onset Diabetes of the Young (HNF1A-MODY). The standard approach for analysing multi-tiered functional datasets has been to evaluate each functional parameter independently. Data from functional characterisation efforts of the HNF-1A protein encoded by the *HNF1A* gene, support that the degree of HNF-1A disruption tends to correlate positively with phenotypic severity: MODY-causing protein-altering variants impair HNF-1A transcriptional activity more severely (≤30% *vs*. wild-type) than *HNF1A* variants associated with increased risk for developing type 2 diabetes in population-specific contexts (40-60% *vs*. wild-type). Rare variants which demonstrated intermediate function (between MODY-casual and wild-type) in transactivation and nuclear localisation assays were shown to be associated with a 6-fold increase in type 2 diabetes predisposition.

*Added value of this study:* We have developed a proof-of-principle analytical framework for robust and unbiased variant stratification using multi-dimensional functional follow-up data from a large number of exome-identified missense variants in *HNF1A*. Through our analytical approach we were able to perform a comprehensive assessment of molecular function by utilising data from as many mechanistic dimensions as possible, avoiding arbitrarily determined cut-offs based on 1D functional data. Our method facilitated informative spatial organization of variants along the *HNF1A* molecular-phenotypic spectrum and an exploration of the contributions of each *in vitro* molecular mechanism on meaningful functional, and therefore clinical, stratification. Further, we were able to perform sensitive mapping of variant effects on molecular function with phenotypic outcome using clinical and genetic data from national MODY diagnostic registries of UK and Norway. This effort allowed us to annotate functional clusters with clinical knowledge and identify discordant classifications between functional genotype and clinical phenotype.

*Implications of all the available evidence:* Our novel approach towards analysing large functional datasets enables sensitive variant-phenotype mapping and multi-layered variant annotation. It also assists in prioritisation of functional elements and signatures for Multiplexed Assays of Variant Effects (MAVEs) whilst they largely remain limited to a single functional readout. Indeed, comprehensively annotated *HNF1A* variant clusters can aid in the interpretation and clinical classification of variants, and can also be utilised to calibrate supervised variant classification models built with high-throughput-derived experimental data.

## Introduction

Precision medicine increasingly relies on an accurate interpretation of the consequence of genetic variation. Large-scale multi-ethnic genetic sequencing studies have challenged our understanding of the relationship between coding variants in Mendelian disease genes, including those involved in monogenic forms of diabetes such as *HNF1A*. Until relatively recently the consensus has been that heterozygous highly penetrant loss-of-function alleles in *HNF1A* give rise to a clinically distinct diabetes subtype, characterised by an early age of onset (typically <25 years), dominant inheritance, sensitivity to sulphonylureas, non-obesity and termed HNF1A-Maturity-onset Diabetes of the Young, (HNF1A-MODY).^1^ Whilst this genotype-phenotype correlation is true for a subset of *HNF1A* variant carriers, it represents one end of a broad spectrum of *HNF1A* variant effects.^2-4^

Genome-wide association and next-generation sequencing studies of randomly ascertained individuals have challenged binary assumptions and overinflated pathogenicity estimates regarding variants in *HNF1A* (and other Mendelian disease genes) and identified common coding variants of low effect associated with increased risk of type 2 diabetes.^5,6-8^ Whole exome sequencing studies in populations of Mexican American ancestry have revealed a low frequency missense variant (p.E508K) in *HNF1A* associated with a five-fold increase in type 2 diabetes prevalence.^3^ These complex genomic insights warrant a more nuanced understanding of the phenotypic manifestation of *HNF1A* gene variants: some alleles are sufficient for early-onset sulfonylurea-responsive diabetes (‘HNF1A-MODY’) in some people, some modify susceptibility for developing complex multifactorial hyperglycemia later in life (type 2 diabetes), and most alleles will likely manifest as benign and neutral.

A correct diabetes diagnosis is important because *HNF1A* is a clinically actionable gene where accurate variant interpretation has tangible impact on diabetes progression and treatment, and implications for family members. Individuals with rare, deleterious *HNF1A* alleles and young-onset diabetes (typically <25 years) are sensitive to treatment with oral sulfonylureas, and can often avoid insulin injections until late in life.^9-10^

The ubiquity of genetic sequencing means that more novel and incidentally detected variants of uncertain clinical significance (VUS) will be identified in individuals with less extreme phenotypes.^11^ The challenge today is in the ability to map *HNF1A* sequence-function relationships at high fidelity, using clinical and molecular characterisation and analytical pipelines with sensitivity to capture the subtleties along the pathophysiological continuum. Rigorous functional follow-up of rare sequence-identified alleles in *HNF1A* is crucial to making correct assignments of pathogenicity. Indeed, functional data are considered a strong line of evidence for accurate clinical diagnostic classification of variants.^12^ Furthermore, it has been shown that diabetes severity in *HNF1A* variant carriers is influenced by allele position in the gene: the transactivation domain is more tolerant to genetic variation and variants in the latter exons^8-10^ are only present in hepatocyte-dominant isoforms and would thus not likely translate to a strong beta-cell phenotype.^13-14^

To understand the relationship between *HNF1A* sequence variation, molecular dysfunction and clinical phenotype, we characterised the functional impact of a total of 73 *HNF1A* missense variants detected in the exomes of 12,940 multi-ethnic type 2 diabetes cases and controls using standard functional assays. Our primary objective was to develop an analytical approach that would enable a) an unbiased and comprehensive evaluation of *HNF1A* variant behaviour based on multiple molecular mechanisms, and b) sensitive mapping of multi-dimensional *in vitro* function to *HNF1A* glycaemic phenotypes *in vivo*. We hypothesised that severity of molecular dysfunction *in vitro* (wild-type/wild-type-like, moderate/intermediate impact, loss-of-function/deleterious) would correlate positively with the severity of clinical phenotype (benign, increased type 2 diabetes risk, young-onset sulfonylurea-responsive hyperglycaemia).

## Methods

The study was approved by the regional ethical committee in Bergen (#2009/2079). We investigated the function of all rare (MAF <0.5%) and low frequency (0.5%<MAF<5%) as well as 3 common (MAF >5%) *HNF1A* nonsynonymous missense variants (n=73) identified in an exome sequencing study of 12,940 type 2 diabetes cases and controls from 5 different ancestry groups^15^ (appendix figure 1; appendix table 1). Collectively, the variants did not enrich for a type 2 diabetes phenotype under any of the several variant filters used (MAF <0.1%, conserved and predicted damaging [PolyPhen: SKAT p=0.30, BURDEN p=0.37]).^15^

### Bioinformatic prediction

The following 4 *in silico* tools were used to evaluate the pathogenicity of the alleles: SIFT^16^, PolyPhen-2^17^, MutationTaster^18^, Combined Annotation Dependent Depletion (CADD).^19^ A CADD cut-off score of 15 was used (>15, pathogenic).

### Functional characterisation

The individual effects of the 73 *HNF1A* missense variants were functionally investigated by 2 research teams at the Universities of Oxford (UK) and Bergen (Norway) using 4 different molecular assays (see detailed description of assays below). Using 2 laboratories allowed us to evaluate the robustness of the functional studies. Each laboratory assessed a unique set of exome-detected variants (n>30), a shared subset of exome-detected variants (n=5), shared type 2 diabetes risk variants (n=2) as well as shared HNF1A-MODY reference variants (positive controls, n=6) (appendix figure 2). The positive controls were selected on the basis of previously reported functional data supporting pathogenicity, clinical evidence for causality (sulfonyurea sensitivity in multiple carriers) and/or genetic (co-segregation) evidence to support their role pathogenesis (appendix table 2). Plasmid and HNF-1A variant constructs, transactivation assays, HNF-1A protein abundance, subcellular localization, and DNA binding are detailed in the appendix.

### Dataset preparation for clustering analysis

The functional datasets were prepared for PCA and clustering analysis by harmonising the number of variables across tested variants. DNA binding ability was only interrogated for a small subset of variants, therefore EMSA data were excluded. Functional data were available in 3 different formats: raw instrument data, data normalised to internal assay controls (renilla luciferase for TA assay, beta-tubulin/actin for protein abundance assay, and nuclear:cytosolic ratio of raw protein abundance reads for nuclear localisation) expressed as biological replicates, and fully processed summary data normalised to wild-type values. The most statistically suitable input format for PCA and unsupervised clustering is functional data normalised to internal assay controls (semi-processed) as intra-assay measurements are harmonised (*vs*. raw instrument data) and the organic structure of the data is retained and uninfluenced by assumptions (*vs*. wild-type normalised data). Further, this format yielded the most robust clustering trends based on distribution quality in multivariate space and known and expected sequence-function relationships. Scores for tissue-specific expression of *HNF1A* isoforms (implications for clinical phenotypic manifestation) and functional domain (varied levels of mutation tolerance) were assigned to each variant (appendix table 3). For stratification of *HNF1A* variants with unsupervised learning methods, see appendix figure 3 and appendix methods.

### Variant-phenotype mapping

We surveyed the UK MODY Diagnostic Registry (Royal Devon and Exeter NHS Foundation Trust, Exeter, UK) and the Norwegian MODY Registry (Haukeland University Hospital, Bergen, Norway) for functionally annotated *HNF1A* missense variants. A total of 162 and 53 *HNF1A* missense variants were documented in the UK and Norwegian diagnostic registries, respectively. Appendix Tables 4 and 5 show the list of clinical features that were available from the database for alleles which overlapped with the Oxford-Bergen dataset (not all features available for each variant). Sequence variants in the Norwegian MODY Registry were classified prior to the incorporation of the ACMG/AMP guidelines^12^, as described^20^ using a 5-tier score system.^21^ Sequence variants in the Exeter MODY Registry had been classified using the ACGS guidelines from 2013 (https://www.acgs.uk.com/quality/best-practice-guidelines/), a 5-tier system used in the UK prior to the advent of the ExAC database and publication of the ACMG/AMP guidelines.^12^ Original clinical reports of carriers were accessed for additional details - particularly where clinical features were sparse - such as extra-pancreatic features, vascular complications, additional family history data, and whether, for example, other MODY genes were next-generation sequenced as part of a MODY gene panel.^22^ The classification system adopted by each centre was used for reclassification of variants from its database.

## Results

### *In silico* & *in vitro* functional characterisation of variants

To resolve *HNF1A* genotype-phenotype complexity we sought to evaluate the function of 73 *HNF1A* missense alleles detected in the exomes of ∼13K multi-ethnic type 2 diabetes cases and controls. The majority of *HNF1A* variants were identified in both type 2 diabetes cases and controls, and for the few observed exclusively in type 2 diabetes cases, they were identified with a frequency of either 1 or 2 cases per variant (appendix table 1). Consensus across multiple *in silico* tools for predicting pathogenicity was observed in only 38 of the 73 variants (53%) (appendix table 1). Based on CADD scores (suggested pathogenicity cut-off >15), ∼70% of the missense variants would be bioinformatically classified as disease-causing (i.e. sufficient to cause MODY).^19^

The 73 *HNF1A* variants were divided between the 2 centres (Oxford and Bergen), and individually evaluated in terms of functional effect using a common pipeline including assays measuring variant effect on HNF-1A transcriptional activity, subcellular localization, protein expression level, and DNA binding ability (appendix figure 1). The Oxford laboratory investigated variants predominantly of South-East Asian aetiology, while the Bergen laboratory studied variants mainly of Caucasian aetiology.

The variants demonstrated a wide range of functional effects from benign to damaging across assays and laboratories with highest variability in transactivation assessments, particularly through regulation of the rat albumin promoter in HeLa cells (activity range 30-110% in Bergen data [appendix figure 4A; 52-114%, Oxford data, appendix figure 5A]). Transcriptional activity was consistently higher for variants using *HNF4A* P2 promoter in INS-1 cells (*vs*. rat albumin promoter in HeLa cells) (activity range 40-90% Bergen data, 77-158 Oxford data), and most likely due to interference of endogenous HNF-1A in INS-1 cells (2-4 fold higher basal promoter activity, appendix figures 4B and 5B). In assessments of protein abundance, >85% of all variants displayed adequate HNF-1A protein levels (>60%, appendix figures 4C and 5C). Similarly, in nuclear translocation assays, most variants were predominantly detected in the nucleus (level >60%), the exception being 5 variants from the Bergen dataset (p.A276D, p.S487N, p.S604R, p.T441K, p.R131Q) (appendix figure 4D) and 5 from the Oxford dataset (p.N62S, p.L518F, p.S535R, p.T537M, p.R583Q) displaying <50% level. The subsets of variants investigated by EMSA demonstrated overall normal DNA binding ability (∼95% of variants >60%), with the exception of one variant from the Bergen dataset (p.R131Q <50%) and 5 from the Oxford dataset (p.N62S, p.R114C, p.T156M, p.A161T, p.S535R; <50%) (appendix figures 4E and 5E).

### Multi-dimensional data analysis

We performed unsupervised stratification of the *HNF1A* variants using the multi-dimensional *in vitro* functional data (semi-processed data normalised to internal technical controls in each assay) supplemented with scores for isoform expression and functional domain, with the aim of mapping molecular dysfunction to clinical phenotype. Each of the 2 datasets were analysed independently to minimise the interference of inter-laboratory variability with true biological signal. We used principal component analysis to facilitate informative dissection and visualisation of multi-parametric functional data. Eigendecomposition of the data matrices revealed transcriptional activity as the greatest contributor to data variance and structure (figure 1). To enable variant subgroup discovery for function-phenotype mapping, data were partitioned using a) k-means clustering in PC space (figure 2) and b) hierarchical clustering using data coordinates from the total number of informative principal components for each dataset (figure 3). The analysis yielded variant clusters neatly organised along the spectrum of *HNF1A* molecular dysfunction ranging from neutral/benign to intermediate to damaging. As such, we broadly annotated the known *HNF1A in vivo* spectrum, from benign to type 2 diabetes risk-modifying to HNF1A-MODY (inherited early-onset hyperglycaemia, likely to be sulfonylurea-responsive based on MODY registry data), onto the principal components plots and dendrograms based on the spatial distribution of the variants along the *in vitro* data-derived functional spectrum, from wild-type/wild-type-like to intermediate to damaging (figures 2 and 3).

**Figure 1.**
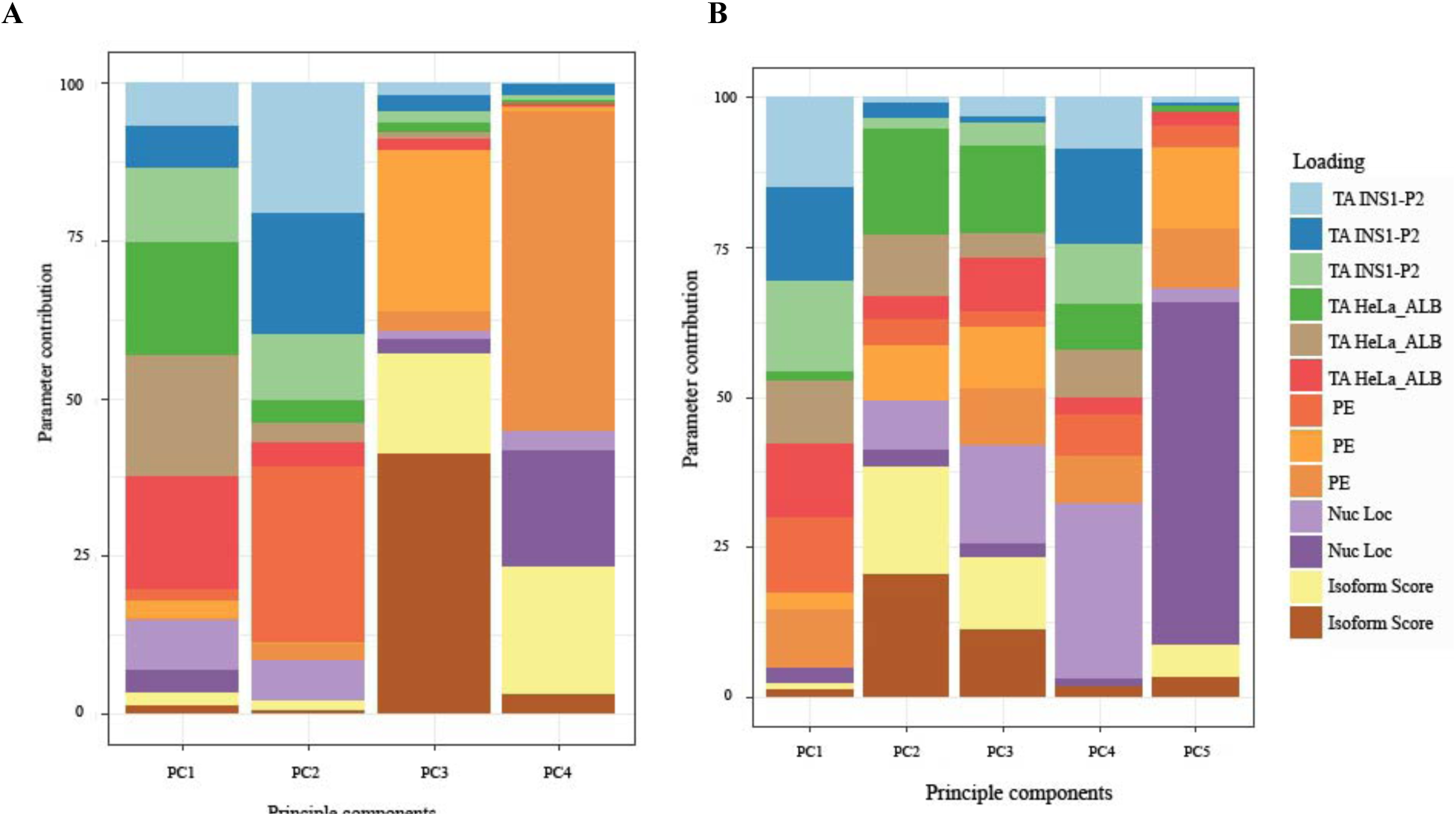
Eigendecomposition of principal components explaining >85% of variance. **(A)** Oxford dataset **(B)** Bergen dataset. TA INS1_P2 and TA HeLa_ALB are transcriptional activity data from INS-1 cells using *HNF4A* P2 promoter and from HeLa cells using rat albumin promoter, respectively. PE = protein expression; nuc loc = nuclear localisation data.

**Figure 2.**
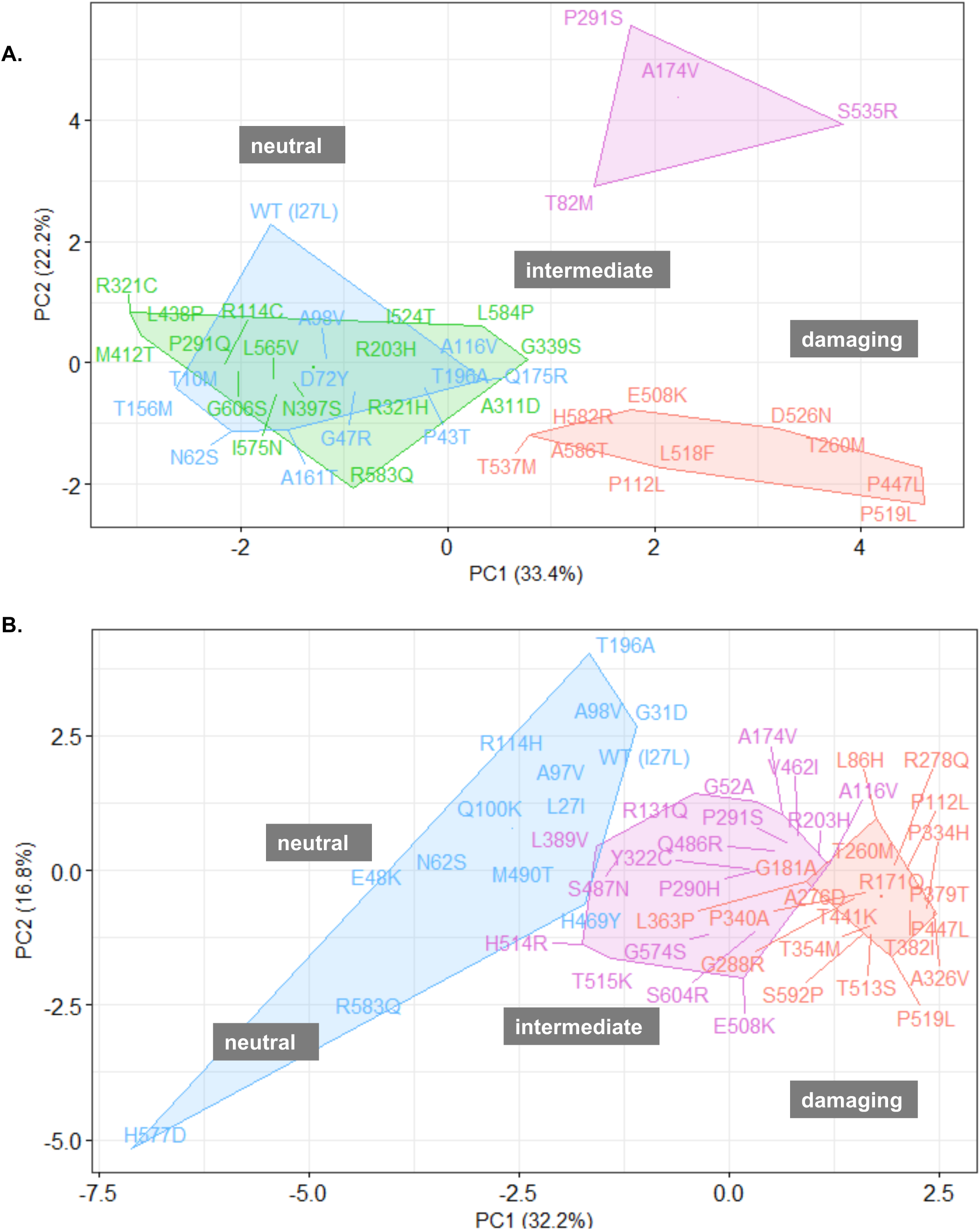
K-means clustering. *HNF1A* missense alleles characterised at Oxford **(A)** and Bergen **(B)** in principal component (PC) space.

**Figure 3.**
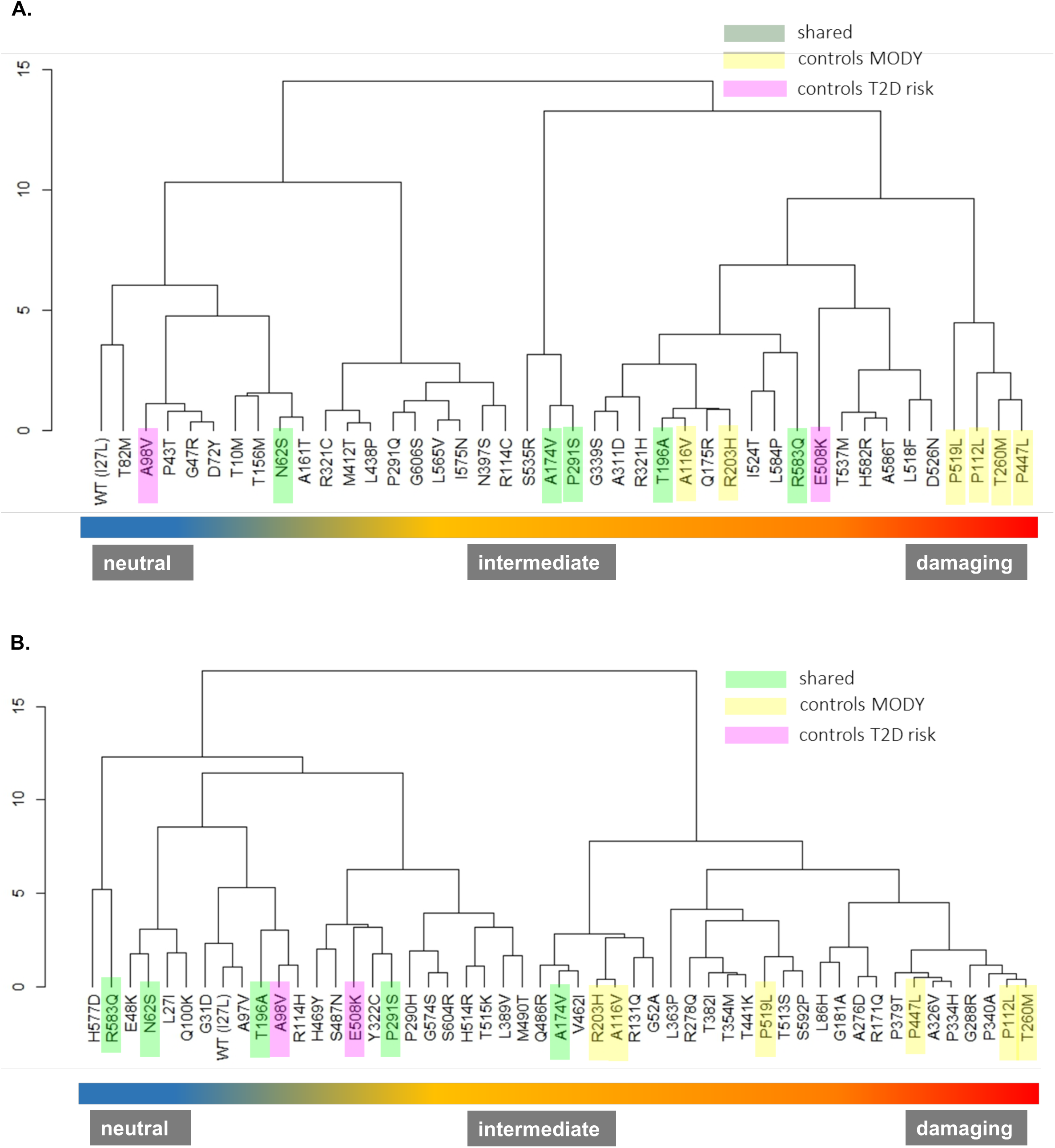
Hierarchical clustering analysis. *HNF1A* missense alleles characterised at Oxford **(A)** and Bergen **(B)**. WARD minimum variance method was used and analysis performed using orthogonally transformed functional data from PC1-PC4 (>85% explained variance) from Oxford dataset and PC1-PC5 (>85% explained variance) form Bergen dataset. To optimise visualisation of the functionphenotype gradient, some branches were rotated.

### Clinical interpretation of *HNF1A* variants

To assess the medical diagnostic utility of multi-tiered *HNF1A* sequence-function annotations, we examined their mapability to *HNF1A* clinical phenotype using clinical data from overlapping *HNF1A* missense variants in the UK and Norway MODY diagnostic registries (appendix tables 4 and 5).

Of the 31 total overlapping variants between our functional effort and the UK registry, 19 were classified as pathogenic/likely pathogenic and 15 as VUS/likely benign. Three of the 31 variants (p.G606S, p.H469Y, p.P291S) were present under both pathogenic/likely pathogenic (where they were considered the MODY-causal variant in the cases) and VUS/likely benign (cases of co-occurrence with a pathogenic variant in *HNF1A* or another MODY gene) classifications. All 15 missense variants categorised as VUS/likely benign in the UK database demonstrated benign clustering patterns in our analysis (i.e. did not form subgroups with variants which exhibited impaired function). However, for 10 of the 19 variants categorised as pathogenic/likely pathogenic in the UK clinical database (p.A161T, p.A174V, p.G47R, p.G606S, p.H469Y, p.M412T, p.N62S, P291S, p.R131Q, p.T10M), patterns of *in vitro* functional clustering patterns did not match clinical diagnostic variant interpretation. The variants either co-occupied clusters with known type 2 diabetes risk modifiers (some moderately impacted in functional assays) or with wild-type/neutral variants. Discordance between functional genotype and clinical variant interpretation prompted a thorough reassessment of variant pathogenicity.

The missense variant p.N62S (GnomAD allele count n=33) was consistently dissimilar to dysfunctional variants in dendrograms and k-means derived clusters from both Bergen and Oxford datasets (figures 2 and 3). In the UK MODY registry, it was identified in an obese individual who was diagnosed with diabetes at age 36 years (appendix table 6). The patient suffered from microvascular complications (nephropathy and retinopathy) (appendix table 6). These features are neither inconsistent with HNF1A-MODY nor a type 2 diabetes phenotype (which might be familial considering the number of affected individuals in the carrier’s pedigree). *HNF1A* was the only MODY gene sequenced in this individual as genetic testing was performed before the advent of the targeted MODY gene exome sequencing panel which is the current diagnostic procedure. Alone, p.N62S allele frequency values are sufficient to confidently re-categorise the variant as VUS/likely benign in the context of MODY (figure 4).

**Figure 4.**
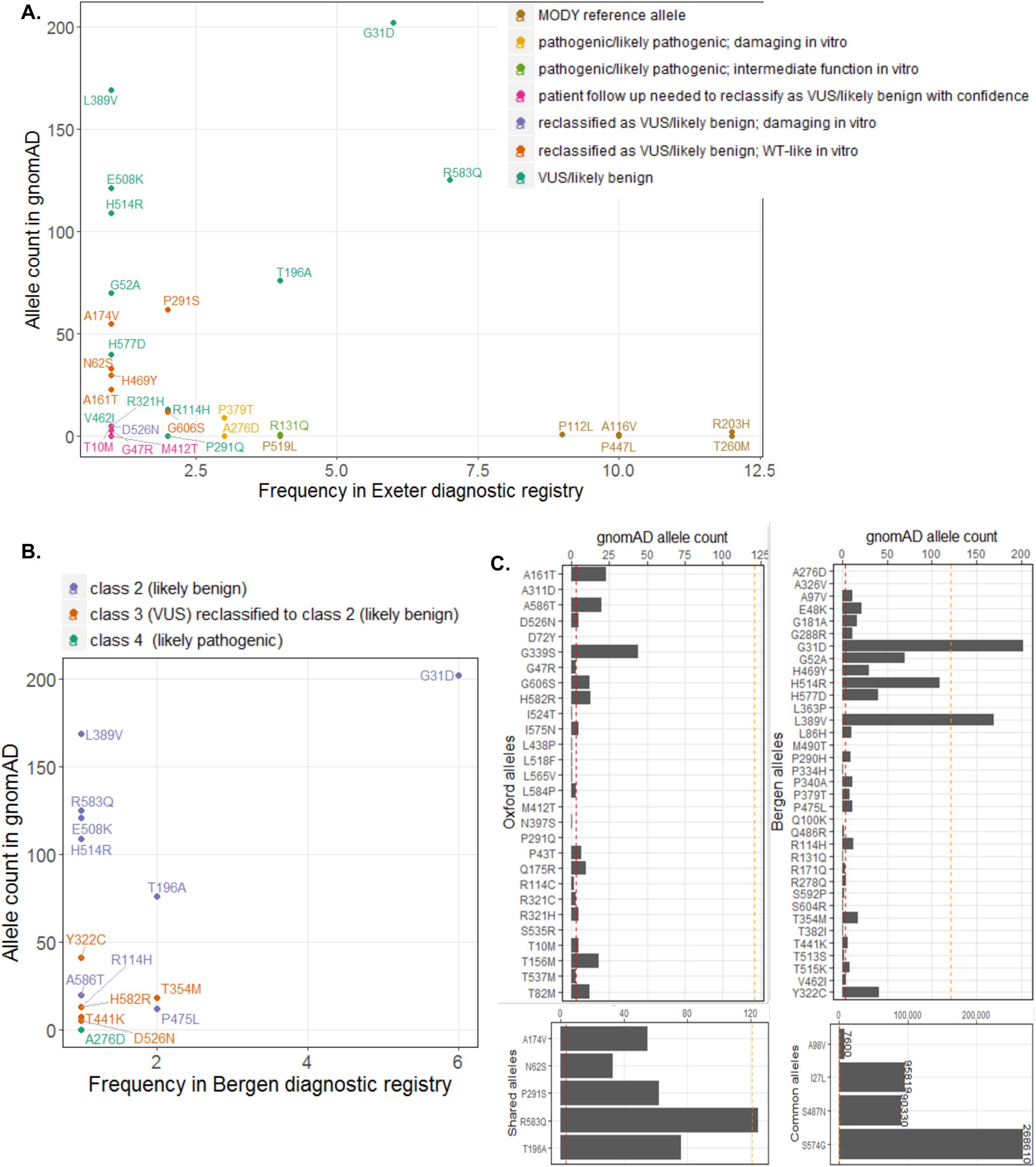
Distribution of functionally annotated *HNF1A* missense alleles. As a function of frequency in the **(A)** UK MODY diagnostic registry and **(B)** Norway MODY diagnostic registry on the x axis and reported frequency in the genome aggregation database (gnomAD) on the y axis. Alleles are coloured on the basis of the (re)classification scheme on the top right. **(C)** Frequency of functionally characterised exome-detected *HNF1A* missense alleles in gnomAD. The red and orange dashed lines mark known ultra-rare, MODY pathogenic (allele count ≤2, AF < 0.0008%) and low frequency type 2 diabetes predisposing allele frequencies (allele count ≤ 121, AF <0.04%) respectively.

The variants p.A174V and p.P291S, characterised by both laboratories, were more difficult to interpret. In the Oxford dataset, these variants formed a separate outlying k-means-derived cluster (figure 2A). They also occupied an independent subgroup in hierarchical clustering which branches high on the height scale away from the larger cluster defined by wild-type and other neutral variants (figure 3A). Atypically high luciferase renilla values (internal luciferase reporter gene assay control used for normalisation) were reported for these variants as well as for p.S535R, which have resulted in a potentially exaggerated reduction in transactivation values for these variants upon normalisation to the internal assay reference in the Oxford dataset (appendix figure 5). In the Bergen dataset, these variants also consistently lie in the type 2 diabetes risk modifier zone (not pathogenic for MODY) (appendix figures 2B and 3B). Not only are the activity profiles of p.A174V and p.P291S dissimilar to those of pathogenic MODY variants, they also occur at a much higher frequency in the general population (figure 4). In terms of clinical profiles, p.A174V was detected in an individual with diet-controlled diabetes, which was diagnosed at age 25 years (appendix table 6). The p.P291S variant was detected in an overweight individual who was diagnosed with diabetes at age 42 years when it was classified as likely pathogenic/pathogenic (appendix table 6). In another unrelated individual, p.P291S was co-expressed with p.G31D and both were annotated as VUS/likely benign in the diagnostic database.

The clinical diagnostic classifications of p.G606S and p.A161T did not match clustering patterns in multivariate space (figures 2A and 3A). The variants did not impact HNF-1A function in the *in vitro* assays tested. The frequency associated with these variants (n=12 alleles in gnomAD and n=2 in the Exeter diagnostic clinic) are inconsistent with those of rare MODY-causing variants. The p.G606S variant has also been found in a single case of hyperinsulinemic hypoglycemia (on diazoxide treatment) in the UK registry and in this case was classified as VUS/likely benign.

In clustering analysis, p.H469Y occupied either the same or highly similar (adjacent) subgroups as wild-type (L27 and I27) (figure 3B). The clinical features of the variant carrier described in appendix table 6 are consistent with severe young-onset familial diabetes however, the allele high frequency in gnomAD is 32. Another variant in *HNF1A*, p.G207D (not present in gnomAD), was, however, detected in the same individual. It was identified in 3 other cases (including co-occurrence with p.H469Y) and was classified as pathogenic/likely pathogenic each time it was identified in the UK MODY Registry. Thus, it is likely that p.G207D is the MODY-causal variant and that p.H469Y is either benign or potentially type 2 diabetes risk-modifying.

Despite alignment between clustering pattern and clinical diagnostic interpretation (figures 2A and 3A), p.D526N was reclassified from pathogenic/likely pathogenic to VUS/likely benign. In transactivation assays HNF1A-p.D526N was the most impaired of all tested exome-identified variants in the Oxford dataset (∼50% in HeLa and ∼80% in INS-1 cells; MODY reference variants exhibited transactivation range of 20-40% in HeLa cells and 30-50% in INS-1 cells in Oxford, with the exception of p.R203H and p.A116V which yielded transactivation values of 40 and ∼60% in HeLa, and ∼50 and ∼100% in INS-1 assays, respectively). The variant is observed only 5 times in gnomAD, which suggests it might not be causal for MODY (figure 4). The clinical profile of the p.D526N carrier did not appear to be consistent with HNF1A-MODY, besides presence of diabetes in 3 generations of the carrier’s family. The variant carrier had BMI 32.4 kg/m2, diagnosed with diabetes at age 33 years. Other clinical features included dyslipidaemia, polycystic ovary syndrome, insulin resistance, and hypertension. The variant was also found in a patient in the Norwegian MODY registry. This patient, diagnosed at 19 years of age, had normal BMI and C-peptide levels. Type 1 diabetes autoantibody status and type 1 diabetes risk score were not known. The carrier was treated with metformin. His mother and the mother’s brother also have diabetes (treated with diet and insulin, respectively). Moreover, the patient was diagnosed with Crohn’s disease. Altogether, this suggests that the carriers might have a combination of type 2 diabetes and HNF1A-MODY, which is not uncommon, or a phenotype representing a possible continuum of diabetes sub-phenotypes from MODY to type 2 diabetes.^5^ Further, the variant is expressed in the hepatocyte-dominant isoform and is thus unlikely to manifest in a strong beta-cell phenotype despite its poor functionality.

Of the 19 *HNF1A* missense variants that overlapped with the Norwegian MODY Registry, 18 were classified as benign (class 1), likely benign (class 2), or VUS (class 3), and 1 (p.A276D) as likely pathogenic (class 4). The variant p.A276D consistently demonstrated impaired HNF-1A function in *in vitro* assays and clustered with the MODY reference variants in the unsupervised clustering analyses, supporting the clinical interpretation of this variant as pathogenic (figures 2B and 3B). It was also classified as likely pathogenic/pathogenic in the UK MODY registry (figure 4A). All variants classified as benign/likely benign/VUS (class 1-3) in the Norwegian registry clustered in the benign or intermediate type 2 diabetes risk modifier zones, with the exception of 4 variants (p.T354M,p.T441K, p.H582R, p.A586T), which demonstrated variable trends across clustering methods (figures 2B and 3B).

In k-means clustering along principal component 1 and principal component 2, these variants co-occupy a hard cluster with MODY reference variants and variants which exhibited damaging *in vitro* function. This is not entirely unexpected for HNF1A-p.T441K which displayed reduced activity (∼50% on both promoters in INS-1 and HeLa cells) and with reduced (<40%) nuclear localization. Although, in hierarchical clustering, where (dis)similarity between variants was determined using principal component scores from all principal components contributing to >85% of overall variance, the trends were more consistent with clinical features and classification; p.T441K and p.T345M are in the type 2 diabetes risk modifier space of the *in vivo* continuum, hierarchically distanced from the subcluster defined by the majority of MODY reference variants and pathogenic damaging variants p.A276D and p.P379T. The clinical phenotypic data of the p.T354M variant carriers seems more consistent with type 1 diabetes, and in the p.T441K variant carrier, another variant in *HNF1A* (p.G292Rfs) was considered the pathogenic MODY variant (appendix table 7). Moreover, the population frequency values of p.T441K (gnomAD allele count n=18) and p.T354M (gnomAD allele count n=7) are slightly higher than expected for rare disease-causing variants (figure 4). As for p.H582R (gnomAD allele count n=14) and p.A586T (gnomAD allele count n=20), in hierarchical clustering, these variants form a subgroup defined by liver isoform variants which demonstrated suboptimal function in one or more *in vitro* assays. Much like p.D526N, these variants are likely to be strong type 2 diabetes risk modifiers. The p.H582R variant carrier was diagnosed with diabetes age 11 years, she had a BMI of 29 at referral one year later, and C-peptide was measured to 1000 pmol/l (appendix table 7). The p.A586T variant carrier was diagnosed at age 11, C-peptide positive (78 pmol/l), GAD, IA2, ZnT8 negative, with no known family history of diabetes, and treated with insulin (appendix table 7).

Based on this comprehensive variant re-assessment effort, we changed the classification of 7 out of 31 variants shared with the UK MODY diagnostic database (p.A161T, p.A174V, p.G606S, p.H469Y, p.N62S, P291S, p.D526N) from likely pathogenic to VUS/likely benign (figure 4A) and all 5 variants categorised as VUS (class 3) or VUS/likely benign (class 3-) in the Norway MODY registry (p.Y322C, p.T354M, p.T441K, p.D526N, and p.H582R) to likely benign (class 2) (figure 4B). This represents ∼23% and ∼26% of total *HNF1A* missense variants in the UK and Norway MODY registries, respectively, that overlap with the functionally interrogated *HNF1A* missense variants detected in the exomes of ∼13K multi-ethnic type 2 diabetes cases and controls.

## Discussion

In this study, we investigated the functional impact of 73 missense variants in *HNF1A*, detected by exome sequencing of a multi-ethnic type 2 diabetes case-control cohort from 4 different mechanistic angles (appendix figures 1 and 2). We present a novel approach for the analysis of multi-parametric functional data which, in the context of *HNF1A*, has enabled i) a holistic assessment of variant behaviour by combining as many mechanistic dimensions as possible, ii) unbiased stratification along the spectrum of glycaemic phenotypes ranging from neutral/benign effects, to modification of multifactorial polygenic diabetes risk, to deleterious and causal for early-onset sulfonylurea-responsive diabetes, iii) an assessment of the relative contributions of each functional parameter to molecular variability, and iv) rigorous phenotype mapping and a thorough re-evaluation of the clinical classifications of overlapping variants in 2 national MODY diagnostic registries

Revisiting clinical variant classifications using *HNF1A* functional clusters led to the reclassification of ∼4% (7/162) and ∼9% (5/53) of all *HNF1A* missense variants in the UK and Norwegian MODY diagnostic registries, respectively. Decisions on variant reclassification were primarily motivated by the juxtaposition of allele frequency values in the general population (based on gnomAD allele counts) against their frequency in the MODY diagnostic registries (highest frequency values belonged to bona fide loss-of-function alleles used as MODY reference controls in this study) (figures 4A and B). Information from other layers of variant annotation such as *in vitro* function (in the tested assays), clinical features, family history, ethnicity, *in silico* prediction all helped to support reclassification decisions.

Dissection of the individual principal components revealed transactivation to be the primary contributor to the spatial distribution of multi-parametric data. This suggests that it might be a superior functional readout and potentially more informative than other molecular assays for assessing *HNF1A* variant pathogenicity, and in line with our previous experience on various functional assays of *HNF1A* variants.^2^ Since transactivation is a relatively all-encompassing measure of transcription factor protein function, we assumed defects in transcriptional activity would capture defects in its biochemical prerequisites (protein expression, nuclear transport, DNA binding). While this may have been the case for the majority of functionally interrogated *HNF1A* variants, p.R203H and p.A116V highlight the limitations of this assumption. For these variants, severely impaired DNA binding ability was not adequately captured by transactivation. This might explain clustering of p.A116V and p.R203H in the intermediate zones amongst type diabetes risk modifiers, away from MODY reference alleles, in both k-means and hierarchical clustering for both centers.

We were able to mitigate error associated with handling data from 2 centers with methodological differences by benchmarking several HNF-1A variants (benign, type 2 diabetes risk, and MODY) in both laboratories. Discrepancies between the 2 centers with respect to shared variants can be explained in part by variability in technical protocols between laboratories and the handling of samples by various individuals over the course of the study. The relative clustering position of the shared variants is impacted by the function trends observed in each dataset; whilst the majority of Oxford variants behaved wild-type-like, the Bergen dataset was more complex as variants demonstrated a wider range of effects. For instance, for an intermediate variant and a known type 2 diabetes risk modifier such as p.E508K, the dissimilarity to MODY reference alleles is more pronounced in the Bergen dataset where there are more data points between moderately impaired and damaging function.

While important, informative and powerful first lines of evidence, functional annotations should not to be treated as superior or stand-alone determinants of variant pathogenicity, which they have not in this study. Indeed, the same variant in a MODY gene can give rise to a spectrum of clinical phenotypes and exhibit variable penetrance depending on genomic (regulatory variants in *cis* or *trans*, or haplotype epistasis) and environmental (epigenomic) context which are difficult to capture in functional assessments.^23-28^ It is also entirely possible that some of the noise in functional-clinical mapping is a reflection of the heterogeneity in the clinical phenotypic manifestation of *HNF1A* variants.^5^ Another aspect to consider is the expected variation in clinical practice between the two centers in the UK and Norway to which diabetes patients have been referred. It would thus be naive to attempt to draw conclusions regarding variant effects *in vivo* from, for example, a single registry observation. The reality of phenotypic variant manifestation is often complex, context-dependent, non-linear, and spectrum-based. Developing a contextual and thorough understanding of variant behaviour from diverse functional, clinical, biochemical and demographic datasets is necessary to facilitate highly accurate interpretation.

The p.D526N variant in *HNF1A* is a perfect example which illustrates the need for nuanced evaluations of variant effects despite the availability of multiple layers of functional annotation from various cell systems. The variant was clinically classified as pathogenic/likely pathogenic and exhibited impaired *in vitro* functional activity (shared a cluster with known MODY-causal variants). Its impact on molecular function would be consistent with biomarker profiles (hsCRP and glycans) suggestive of HNF1A-MODY. Yet, re-evaluation of the clinical features of variant carriers in UK and Norway diabetes registries, and the fact that it is present only in the longest *HNF1A* transcript isoform expressed predominantly in liver, suggest that it is more likely to be a contributing factor to common multifactorial diabetes rather than a primary driver of early-onset sulfonylurea-responsive familial hyperglycaemia. Integration of isoform weights into the unsupervised clustering model helped separate bona fide loss-of-function MODY variants from functional variants expressed in exons 8-10 at the lowest level of hierarchical clustering.

The recent advent of Multiplexed Assays of Variant Effects (MAVEs) has made it possible to interrogate the function of every possible sequence perturbation in a single experimental system.^28^ Successful and productive implementation of these technologies requires overcoming the technical and analytical complexities associated with scaling up, which represent the most significant barrier in the face of closing the chasm between variant resolution and variant interpretation. A meticulously designed MAVE for *HNF1A* would enable functional annotation of all possible missense variants (>12,000) in a single assay. A high-performance variant classifier built using MAVE-based data can then be used to generate an exhaustive catalogue of variant effects which researchers and clinicians can consult upon sequence-identification of an *HNF1A* variant.

In conclusion, we have developed a novel analytical framework for robust and unbiased variant stratification using multi-dimensional functional follow-up data from the largest number of exome-identified missense variants in *HNF1A* ever studied. This allowed us to annotate functional clusters with clinical knowledge and identify discordant classifications between functional genotype and clinical phenotype. We believe our pipeline is an important proof-of-principle technical contribution on the path towards more reliable, scalable, and comprehensive mapping of sequence-function relationships: a significant factor in making well-informed initial judgements of allele pathogenicity on the context of individual phenotypic presentations.

## Data Availability

Summary statistics is available for all data in the article.

## Contributors

SA, LB, PRN, and ALG were responsible for the concept and design of the study. SA, LAN, JKR, AJB, KC, IA, AM, SE, MIM, LB, PRN, and ALG were involved in, protocol development, data collection, data interpretation, and critically revising the report. NH, SN, AK, TVDL, and JM were involved in data collection, data interpretation, and critically revising the report. All authors reviewed and approved the manuscript for submission.

## Acknowledgements

ALG is a Wellcome Senior Fellow in Basic Biomedical Science. SE and MIM are Wellcome Senior Investigators. This work was funded in Oxford by the Wellcome (095101 [ALG], 200837 [ALG], 098381 [MIM], 106130 [ALG, MIM], 203141 (ALG, MIM], 203141 [MIM]), Medical Research Council (MR/L020149/1) [MIM, ALG], European Union Horizon 2020 Programme (T2D Systems) [ALG], and NIH (U01-DK105535; U01-DK085545) [MIM, ALG]. The research was funded by the National Institute for Health Research (NIHR) Oxford Biomedical Research Centre (BRC) [ALG, MIM]. This work was funded in Bergen by grants from the European Research Council (#293574 [PRN]), the Norwegian Research Council (#240413/F20 [PRN]), Stiftelsen Kristian Gerhard Jebsen [PRN], the Novo Nordisk Fonden [PRN], the Western Norway Health Authorities, (#911745 [PRN]), and the University of Bergen [IA, LN, PRN].

